# Associations between weather and *Plasmodium vivax* malaria in an Amazonian elimination setting: a distributed lag analysis from 2017–2024

**DOI:** 10.1101/2024.11.26.24318000

**Authors:** Gabriella Barratt Heitmann, Xue Wu, Anna T. Nguyen, Astrid Altamirano-Quiroz, Sydney R. Fine, Bryan Fernandez-Camacho, Antony Barja, Renato Cava, Verónica Soto-Calle, Hugo Rodriguez, Gabriel Carrasco-Escobar, Adam Bennett, Alejandro Llanos-Cuentas, Erin A. Mordecai, Michelle S. Hsiang, Jade Benjamin-Chung

## Abstract

**Background:** *Plasmodium vivax* (*Pv*) is the predominant malaria species in countries approaching elimination. Environmental factors can guide intervention targeting.Yet, research that considers the long-term relapse periodicity of *Pv* is limited, particularly in Latin America.

**Methods:** We merged *Pv* malaria incidence data from 2017–2024 from 136 communities in the Peruvian Amazon with hourly weather data from the ERA5 dataset. Predictors included weekly minimum and maximum temperature and total weekly precipitation. We fit non-linear distributed lag models using a lookback period of 2–16 weeks. We conducted sub-group analyses by community type (adjacent to river versus highway) and El Niño Southern Oscillation (ENSO) period. Temperature models were adjusted for total precipitation; precipitation models were adjusted for maximum temperature.

**Findings:** Minimum temperature at the 90th percentile (23.7°C) was associated with 10% (95% CI 5%–14%) higher malaria incidence compared to the lowest minimum temperature (20.4°C) at a 7-week lag. Maximum temperature at the 90th percentile (33.7°C) was associated with 10% (95% CI 8%–13%) higher malaria incidence compared to the lowest maximum temperature (29.6°C) at a 9-week lag. Total weekly precipitation at the 90th percentile (1000mm) was associated with 29% (95% CI 24%–33%) higher malaria incidence compared to weeks with no precipitation at an 11-week lag. Incidence was higher and associations were stronger in communities adjacent to rivers versus highways. Malaria incidence was lower during El Niño periods, and there was evidence of interaction on the multiplicative scale for the association between incidence, all weather predictors, and ENSO period.

**Interpretation:** *Pv* malaria incidence was positively associated with higher temperatures and precipitation in an elimination setting in Peru, particularly in riverine communities during non-El Niño years, with longer lag periods than previously reported for such associations. These findings can inform malaria elimination interventions to combat the long-lasting effects of weather on *Pv* transmission.

**Research in context:** *Evidence before this study:* We searched PubMed for (“weather” OR “climate” OR “temperature” OR “precipitation“) AND (“malaria“) AND (“Amazon“). The search yielded 76 results. The most relevant studies looked at the influence of weather variables on all malaria incidence in neighboring Brazil and Venezuela. The study in Brazil found positive associations with temperatures between 25–30°C and precipitation >4.46cm at 1-week lags, but negative associations with temperatures >25°C at 2–3-week lags. The study in Venezuela found positive associations with mean temperatures in the range 20–30°C and no association with precipitation. Both studies emphasized that malaria transmission was highly heterogeneous.

*Added value of this study:* Our study used a distributed lag analysis to account for the long-term relapse periodicity common in *Pv* malaria, which is predominant in the Peruvian Amazon. Our study is, to our knowledge, the first to exclusively focus on climatic drivers of *Pv* malaria at long-term lags in an elimination setting dominated by the *Ny. darlingi* vector. This transmission system is especially understudied compared to *Plasmodium falciparum* malaria, yet *Pv* malaria is the predominant species in elimination settings worldwide.

*Implications of all the available evidence:* Our study contributes to an important body of literature characterizing climatic drivers of *Pv* malaria in the Peruvian Amazon. Our findings indicate that over 1–4-month lags, higher temperatures and precipitation could contribute to higher malaria incidence, particularly during neutral El Niño Southern Oscillation (ENSO) years. We also found that communities near rivers were more sensitive to changes in temperature, while communities near highways were more sensitive to changes in precipitation.

## INTRODUCTION

As countries approach malaria elimination, the proportion of malaria infections due to *Plasmodium vivax* (*Pv*) increases.^1^ *Pv* presents unique challenges for malaria elimination efforts because of its ability to survive in a wide range of environments, including temperate settings, by utilizing a dormant hypnozoite stage.^2^ Relapsing infections occur weeks to months after a prior infection or relapse, and serve as a significant reservoir for persistent transmission.^3^ ^4,5^ *Pv* infections can be latent, as well asymptomatic, sub-patent, or minimally symptomatic, making diagnosis and surveillance a challenge. Further, symptomatic infections are low density and often missed through standard diagnostics.^4,6,7^ These characteristics greatly obfuscate *Pv* transmission biology and make *Pv* harder to detect, treat, and ultimately eliminate.

Over 90% of Latin America’s malaria burden is concentrated in the Amazon Basin, and and a recent review of malaria elimination progress highlighted the need to study risk heterogeneity within the Amazon.^8^ Parasite populations and vector behavior vary significantly within this region,^9–11^ and this variation is likely driven by adaptations to different environmental conditions.^10,12,13^ The Amazon Basin is also undergoing rapid environmental and climatic changes^14–16^, and malaria elimination will require a fine-scale understanding of how those changes are altering transmission dynamics.

There is strong biologic plausibility for temperature and precipitation to influence *Pv* transmission (Figure 1).^6,17–22^ Increased precipitation can increase standing water presence, soil moisture, and humidity, which in turn increase breeding habitat for malaria vectors.^23^ However, this relationship is complicated: in dry-land communities, heavy rain can flush mosquito larvae, leading to decreased mosquito populations, whereas in riverine settings such as in the Amazon, flooding from heavy rains can increase breeding ground and mosquito populations.^23^ Temperature has strong effects on the capacity of the *Nyssorhynchus* vector to transmit malaria via the mosquito biting rate, mosquito abundance, parasite incubation rate, and mosquito longevity.^24,25^ Finally, weather could be a direct trigger for relapse infections for *Pv* infections^26^, and there is other indirect evidence that relapses may be triggered by antigens in uninfected mosquito saliva (biting rate is temperature-sensitive).^3^

**Figure 1.**
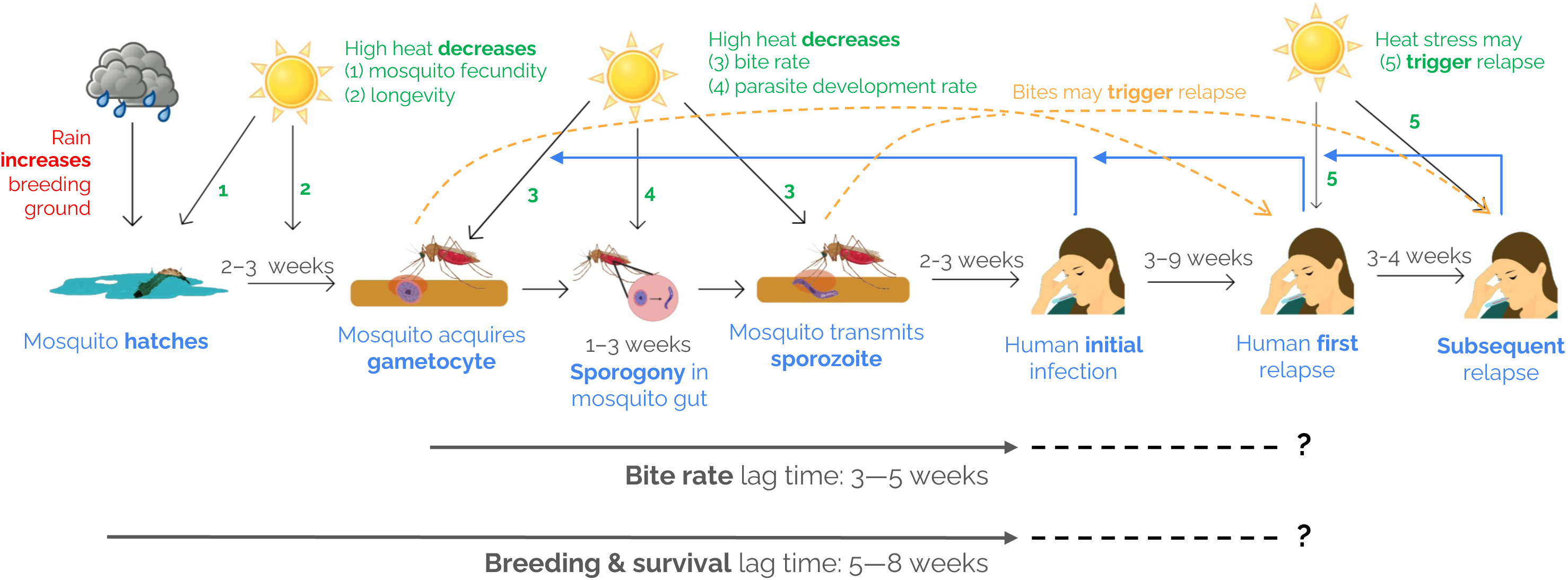

Prior research on the relationship between weather and malaria incidence in Latin America has found positive associations with rainfall but mixed associations with higher temperatures, and there is significant spatial heterogeneity^27,28^. Additionally, prior studies have not focused specifically on *Pv* malaria, including those conducted in the Amazon, where *Pv* accounts for 77% of the transmission burden.^11^10/21/2025 6:09:00 PM The Loreto Region has maximum temperatures between 30–35°C thought to be above the malaria transmission thermal optimum (25°C), and daily temperature extremes are thought to impact malaria transmission^29^. To our knowledge, the thermal biology of *Pv* transmission by *Nyssorhynchus* (formerly *Anopheles*) *darlingi,* the primary vector in this region, has not been well characterized in disease ecology studies.

Furthermore, the Amazon region is subject to longer-term climate oscillations due to the El Niño Southern Oscillation (ENSO). The ENSO cycle is characterized by hot, dry conditions during El Niño years and rainy conditions during La Niña years; these periods oscillate with neutral periods every 3–7 years^15^. One prior study in the Brazilian Amazon found that El Niño and La Niña were associated with decreased malaria incidence, but there was significant spatial heterogeneity^28^.

Our objective was to assess the influence of rainfall and temperature on *Pv* incidence in Loreto Region, Peru, one of the only remaining regions with malaria transmission in the country and key to the Peruvian government’s target of malaria elimination by 2030.^30^ Information about climatic drivers of *Pv* transmission is needed to inform effective tailoring and targeting of interventions for malaria elimination.

## METHODS

### Study design

We analyzed malaria incidence data collected from January 2017 to December 2024 in all 136 communities in San Juan Bautista, Punchana, Alto Nanay, and Belen districts in Loreto region, Peru, an area covering 2,569 km^2^ (Figure 2). The Peruvian Ministry of Health (MINSA) conducted a census in 2017 that was used to estimate the starting population size of each community. The average population size of each community was 321 individuals (range: 15 to 3223).

**Figure 2.**
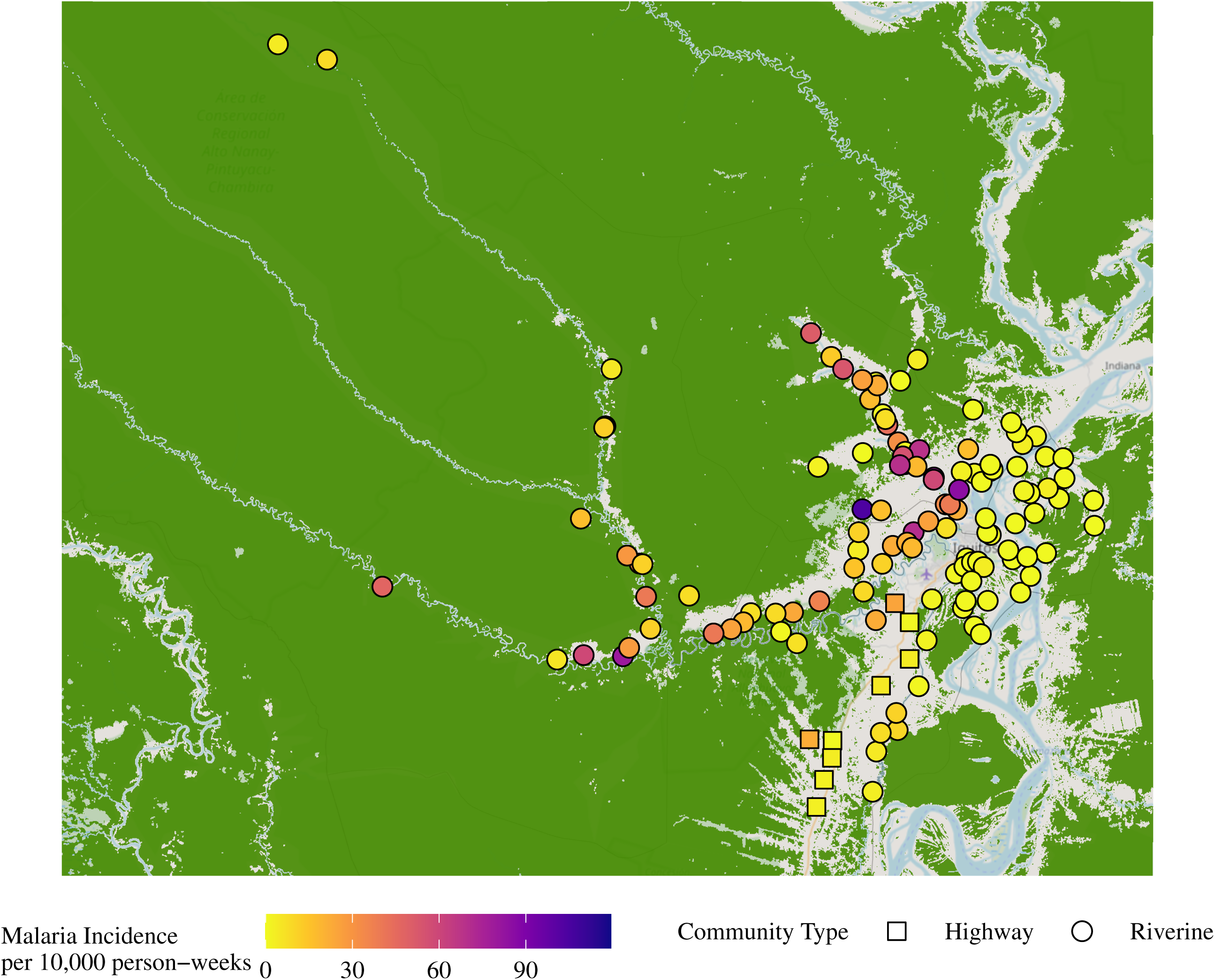

Peru’s malaria burden is concentrated in the Loreto Region, which includes a large swath of the Peruvian Amazon.^7^ Since the 1990s, rapid urbanization and deforestation in the area surrounding Iquitos, the capital of the Department of Loreto, has contributed to increased malaria burden^7^, likely through increased forest edge habitat, which promotes mosquito breeding, survival, and biting^14^. While this region is approaching elimination, sustained, endemic transmission remains, with an annual incidence rate of 17.4 cases per 1000 in 2019,^30^ and *Pv* malaria accounts for approximately 80% of the malaria burden.^7^ Transmission typically peaks in Loreto between February and July^7^ and there is a sizable burden of asymptomatic infections.^18^ Asymptomatic infections typically go undetected and uncounted in routine malaria surveillance in Peru,^18^ since only febrile individuals are tested.

From 2005-2010, there was a large effort towards malaria elimination in Peru via the Project for Malaria Control in Andean Border Areas (PAMAFRO) program that improved case management, including through deployment of community case workers and distribution of insecticide-treated bed nets.^7^ Transmission declined during the project, but the project did not achieve elimination, and transmission has since risen.^31^ Following PAMAFRO, the government of Peru adopted Plan Malaria Cero (PMC) in 2017 to achieve malaria elimination by 2030. Control activities over the study period involved active case detection (ACD), reactive case detection (RACD), distribution of long-lasting insecticide-treated bed nets (ITN), and indoor residual spraying^32^.

### Outcome data

The primary outcome was weekly community-level *Pv* malaria incidence primarily measured through passive case detection conducted by MINSA, though PMC activities including ACD and RACD also occurred over the study period and were recorded in surveillance data. Both physical records stored in health posts and online records compiled by MINSA were used in this analysis. Febrile patients presented for care at a health post and were tested for malaria via microscopy. According to PMC policy^32^, blood smears are stained with 2% Giemsa for 30 minutes. Parasite densities are calculated from the number of asexual parasites per 200 leukocytes (or per 500, if <10 asexual parasites/200 leukocytes), assuming a leukocyte count of 8,000/µL. A blood smear is considered negative if examination of 100 high power fields does not reveal asexual parasites. Thin smears are used for parasite species identification. Slides are read by two microscopists. If there are discordant results, the results are determined by a third microscopist. Only *Pv* cases were included in the present study, and the data did not distinguish between primary infections versus relapse cases. Cases were matched to community census data to confirm community of residence and then matched to geocoordinates for the community centroid. Population counts per community were calculated from a government census conducted from 2017. The FocaL Mass Drug Administration for Vivax Malaria Elimination (FLAME)^33^ trial in Peru (NCT05690841) conducted a census on 30 of these communities in 2023, which was used to estimate an annual population growth rate for each year from 2017–2024. Adjusted population counts per year were estimated by calculating the annual growth rate using the 2023 and 2017 census counts. The same flat annual growth rate was applied to all 136 communities with a population greater than 15 people in the study area.

We obtained data for ITN distribution from MINSA with dates of delivery for communities included in the study from 2018–2024. Time since most recent ITN delivery was discretized into never received, 0–1 year, 1–3 years, and more than 3 years.

### Environmental variables

We obtained temperature and precipitation data from the ERA5-Land Hourly dataset, 11 kilometer resolution, collected by the Copernicus Climate Change Service^34^. We used the air temperature at 2 meters above the surface (K) band for all temperature variables and the total precipitation (m) band for precipitation variables. We matched weather variables to incidence data via community centroid.

We imputed temperature or precipitation values less than 0 as missing. We aggregated hourly temperature values into the weekly minimum and maximum temperature; and total precipitation observed each week. All data extraction was performed using the Python API for Google Earth Engine on Google Colab servers. Aggregation over time and statistical modeling were performed in R version 4.2.1.

We obtained data on the Oceanic Niño Index (ONI), a measure of the strength of the El Niño-Southern Oscillation (ENSO), from the National Oceanic and Atmospheric Association National Weather Service Climate Prediction Center. A 1-unit increase in ONI represents a 1°C higher average sea-surface temperature compared to a 30-year reference period^35^.

We manually designated riverine versus highway communities via Google Earth hybrid satellite image using QGIS version 3.32; we checked our designations with ground-truth observations from 2023 for a subset of communities.

### Statistical analysis

We published a pre-analysis plan at https://osf.io/f89te. Deviations from the pre-analysis plan are listed in Table S1. We removed years 2020 and 2021 from our primary analysis because of strong changes in incidence and case detection resulting from behavior changes and health system capacity during the COVID-19 pandemic.

To assess the timing of the association between malaria incidence and weather variables, we fit distributed lag models. We chose a 2–16-week lookback period (i.e., an infection during the course of week 20 looked back to week 4, 5, …, 18) to account for variation in the potential cumulative effect of rainfall and temperature on *Pv* transmission and long-lasting impacts on relapse cases (Figure 1). For weather exposures (weekly minimum temperature, weekly maximum temperature, and total precipitation), we fit non-linear distributed lag (DL) models^36^ with a log link and Poisson family using weekly malaria case counts per community as the dependent variable with an offset for log community population size using the R package DLNM version 2.4.7^37^. For the distributed lag cross-basis function, we specified a log function with 2 knots to allow for more flexible variation in the short term, and natural splines with 2 degrees of freedom for the non-linear predictors, which yielded the lowest AIC and BIC in model testing. All incidence ratios were calculated relative to reference weeks. In all models, we restricted weather measurements in the input data to observations between the 5^th^ and 95^th^ percentile of the variable’s empirical distribution to remove outlier effects. For temperature predictors, we used 20.5 for minimum temperature and 28.1°C for maximum temperature as references. These values were selected because they correspond to the minimum value within the data range during the study period. For total precipitation, weeks with 0 mm of precipitation were used as the reference. Temperature variables were adjusted for the cross-basis matrix for total precipitation, and total precipitation was adjusted for the cross-basis matrix for minimum temperature, in accordance with AIC/BIC model testing. All DL models included fixed effects for year, community, and years since ITN distribution, and a smooth term for ONI with 3 knots. All positive cases were matched to weather data. Thus, we performed a complete case analysis. Because incidence data for the weather models was aggregated at the community level, we did not test for spatial autocorrelation or adjust for spatial clustering.

To assess the relationship between each predictor and malaria incidence, we fit generalized additive models (GAMs) with a Poisson family and log link and an offset for community population size. We fit models for the week with the strongest association in the DL model (primary lag). All GAMs included fixed effects for year, community, and years since ITN delivery and a smooth term for ONI with 3 knots. We calculated confidence intervals using 1,000 bootstrap iterations with replacement.

We conducted subgroup analyses using the primary lag for temperature and precipitation predictors stratifying by ENSO periods and community type. We defined periods of 5 or more consecutive months of sea surface temperature abnormality of 0.5°C or above/-0.5°C or below as El Niño/La Niña periods, respectively^35^. To assess interaction effects between weather and ENSO period or community type on the multiplicative scale, we fit generalized linear models (GLMs) that included the same covariates from GAM models as fixed effects.

This study was approved by the Institutional Review Board of Stanford University (72291) and by the Dirección Universitaria de Asuntos Regulatorios de la Investigación de la Universidad Peruana Cayetano Heredia (211747).

### Role of the funding source

We received funding from the National Institute of Allergy and Infectious Disease, GSK, and the King Center on Global Development. Funding sources did not contribute to the study design, data collection, analysis, writing, or decision to submit for publication. MH and JBC are Chan-Zuckerberg Biohub Investigators.

## RESULTS

During the study period from January 2017 to December 2024, minimum temperature varied from 17–25°C, and maximum temperature varied from 28–41°C (Figure 3A). Total weekly precipitation varied from 7mm per week to 2,587mm per week (Figure 3B). There were 104 El Niño weeks and 92 La Niña weeks over the study duration, and incidence varied by ENSO period, with the highest incidence during neutral periods. Malaria incidence was highest from 2017–2019, then decreased dramatically in 2020 and 2021 and increased to levels below the 2019 levels in 2022–2024. Malaria incidence did not appear to follow a distinct seasonal trend (Figure 3C). The mean incidence for all communities was 10.4 cases per 10,000 person-weeks.

**Figure 3.**
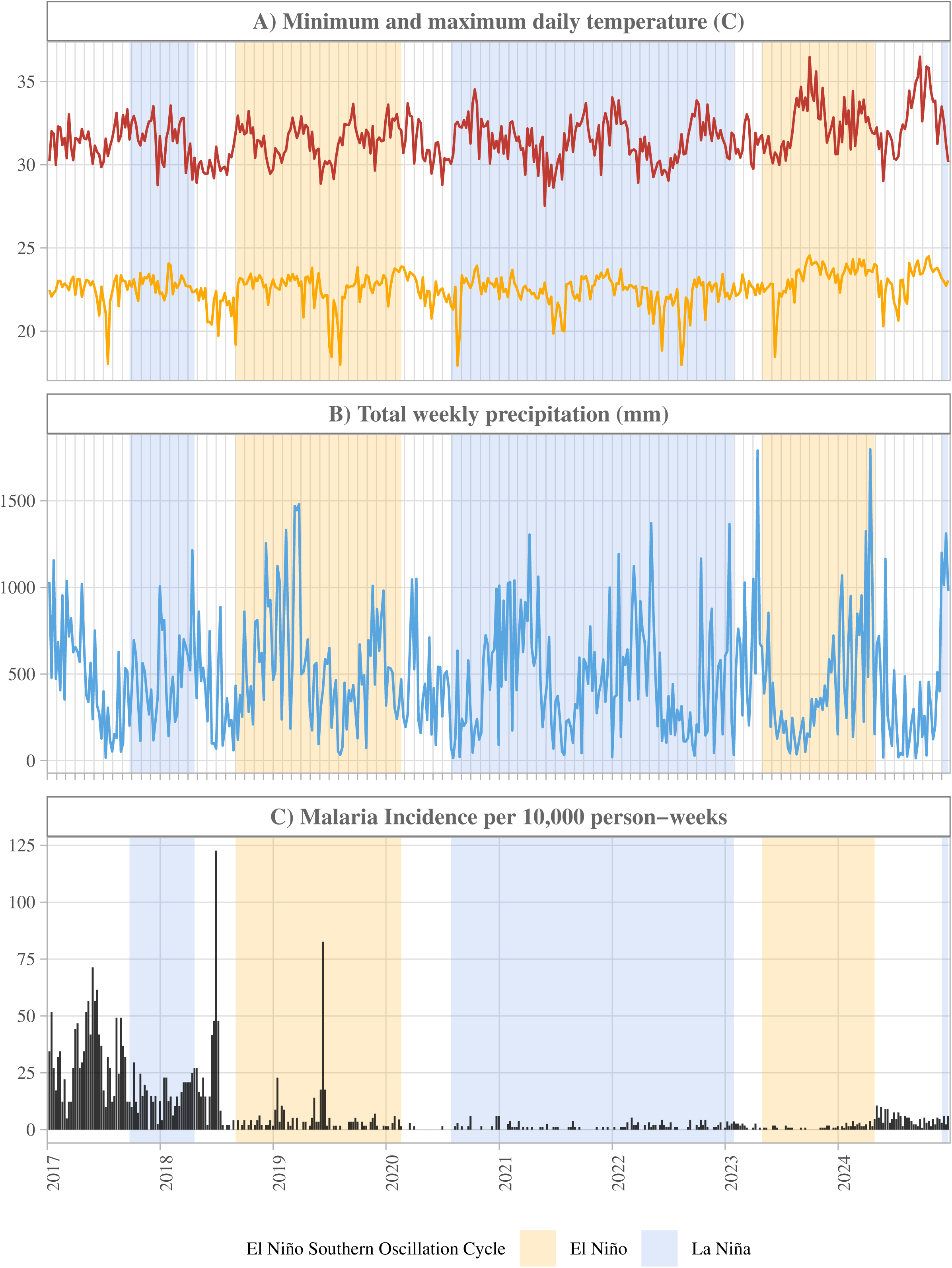

### Temperature

There was a positive association between minimum temperature and malaria incidence at 2–13 weeks lags, and a negative association at 14–16-week lags when comparing the 90^th^ percentile of minimum temperature (23.7°C) to the lowest value (20.5°C) (Figure 4A, 4C). Where this association was highest, at a 7-week lag, the association between minimum temperature and incidence was examined in a marginal effects plot (Figure 4B). At minimum temperatures of 20.5–22°C, incidences were similar. At higher temperatures, there was a positive, non-linear association with incidence of 23 cases per 10,000 at 20.5°C compared to 37 cases per 10,000 at 23.8°C. At a 7-week lag, compared to weeks with the lowest minimum temperature, weeks with a minimum temperature of the 90^th^ percentile were associated with up to 9% higher malaria incidence (95% CI 5%–14%), or incidence ratio of 1.09 (Figure A).

**Figure 4.**
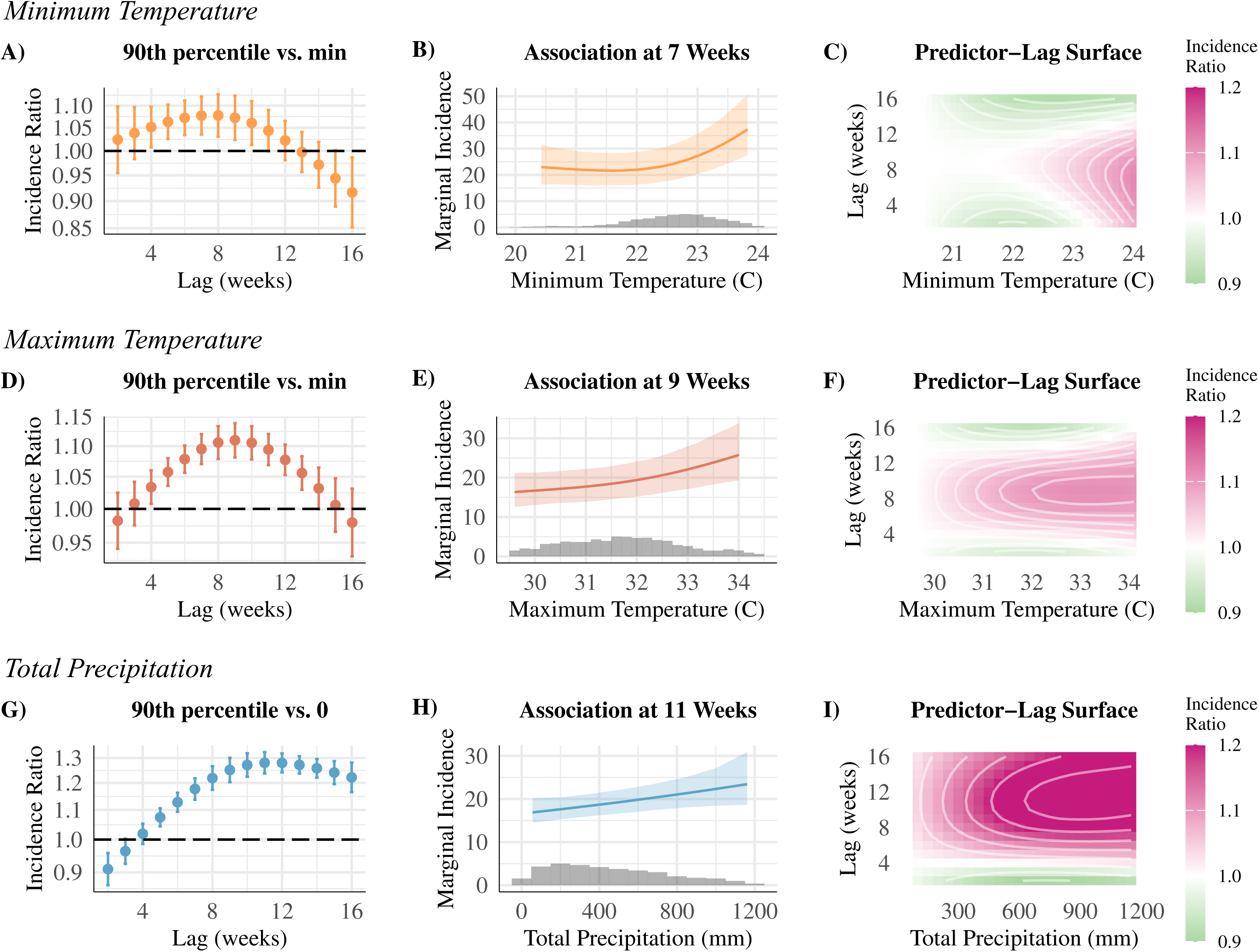

The association between maximum temperature and malaria incidence was non-linear and hump-shaped across lag time; there was a positive association between maximum temperature and malaria incidence at 4–13-week lags when comparing the 90^th^ percentile of maximum temperature (33.7°C) to the lowest value (28.1°C) (Figure 4D, 4F). Where this association was highest, at a 9-week lag, the association between maximum temperature and incidence was examined in a marginal effects plot (Figure 4E). Incidence increased from 16 cases per 10,000 to 26 cases per 10,000 as maximum temperature increased from 29.6°C to 34°C. Compared to weeks with the lowest maximum temperature, weeks with a maximum temperature of the 90^th^ percentile were weakly associated with up to 11% higher malaria incidence (95% CI 8%–14%), or incidence ratio of 1.11 at a 9-week lag (Figure 4D).

### Precipitation

The association between precipitation and malaria incidence was non-linear and positive at lags of 5–16 weeks, when comparing the 90^th^ percentile of precipitation (1000mm) to a reference of no precipitation (Figure 4G, 4I). Where this association was highest, at a 11-week lag, the association between maximum temperature and incidence was examined in a marginal effects plot (Figure 4H). Incidence increased from about 17 cases per 10,000 to about 23 cases per 10,000 as precipitation increased from 50mm to 1150mm (Figure 4H). At an 11-week lag, compared to weeks with no precipitation, weeks with precipitation at the 90th percentile were associated with up to 28% higher malaria incidence (95 CI 24%–32%), or incidence ratio of 1.28 (Figure 4G).

### Community Type Sub-group

Malaria incidence was higher in riverine versus highway-adjacent communities (Table 1, Figure 5A–C). Using GLMs, the association between malaria and minimum temperature was similar by community type, and there was no evidence of interaction on the multiplicative scale (Table 1). However, when using GAMs that allowed for non-linear relationships, the association with incidence and minimum temperature was stronger in riverine versus highway-adjacent communities at minimum temperatures above 22°C (Figure 5A). For maximum temperature, GLMs showed associations were stronger in riverine versus highway-adjacent communities: the incidence ratio for a 1°C increase in maximum temperature was 1.13 (95% CI 1.11–1.15) in riverine communities compared to 1.03 (0.98–1.09) in highway communities (interaction *p* = 0.001) (Table 1). For precipitation, GLMs showed associations were weaker for riverine versus highway-adjacent communities: the incidence ratio for a 500mm increase in precipitation was 1.06 (95% CI 1.02–1.09) in riverine communities compared to 1.58 (95% CI 1.44–1.74) in highway communities (interaction *p* < 0.001) (Table 1).

**Figure 5.**
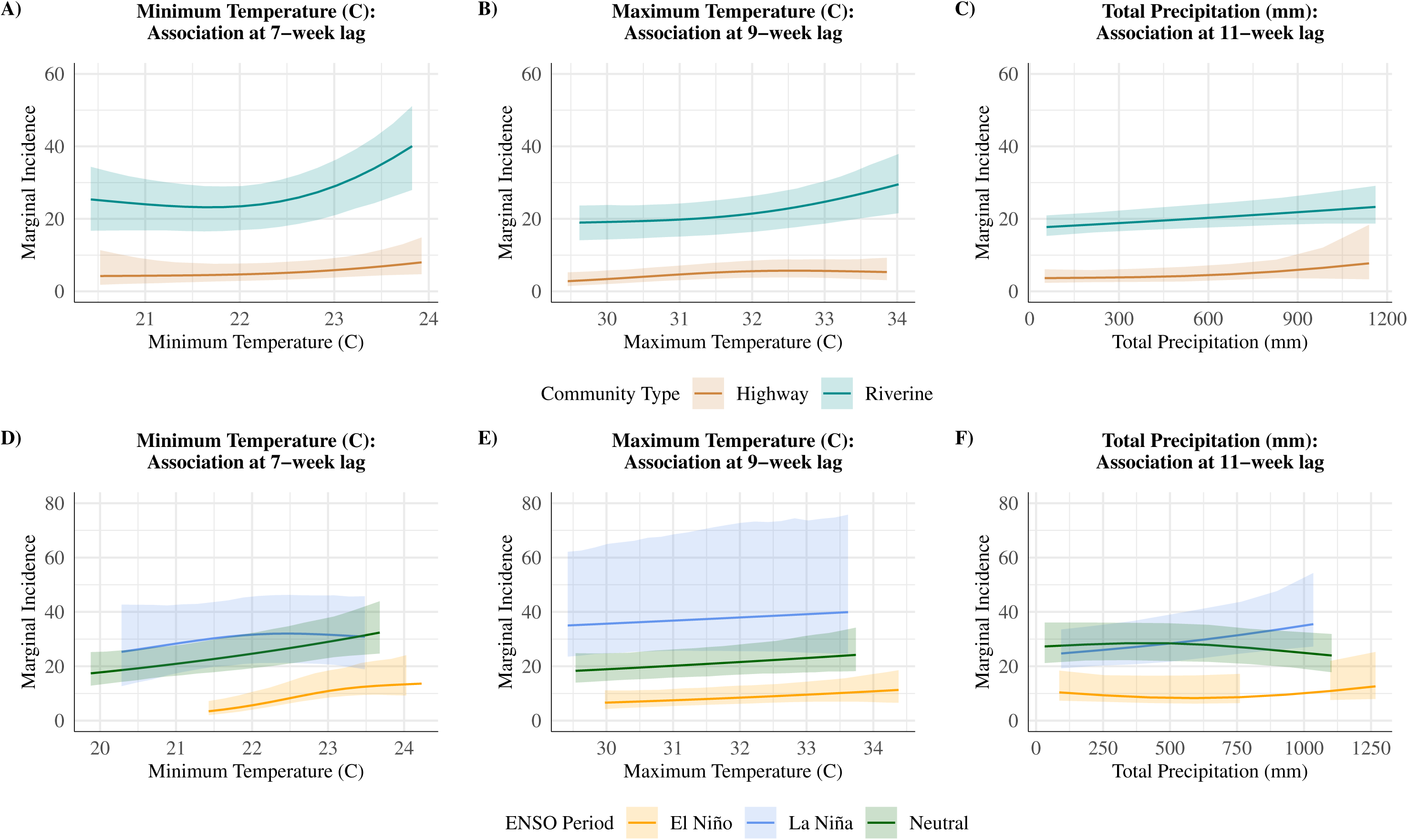

**Table 1.**

| Community type | Predictor range | Incidence range | Incidence Ratio (95% CI) | Interaction p-value |
| --- | --- | --- | --- | --- |
| **Minimum temperature** |  |  |  |  |
| Highway | 20.5–23.9 | 4.2–8.0 | 1.17 (1.08–1.28) | -- |
| Riverine | 20.4–23.8 | 23.2–40.0 | 1.27 (1.23–1.31) | 0.097 |
| **Maximum temperature** |  |  |  |  |
| Highway | 29.5–33.9 | 2.8–5.7 | 1.03 (0.98–1.09) | -- |
| Riverine | 29.6–34.0 | 19.0–29.5 | 1.13 (1.11–1.15) | 0.001 |
| **Precipitation** |  |  |  |  |
| Highway | 51–1139 | 3.6–7.7 | 1.58 (1.44–1.74) | -- |
| Riverine | 57–1161 | 17.7–23.3 | 1.06 (1.02–1.09) | <0.001 |

### ENSO Sub-group

Malaria incidence was lower during El Niño periods compared to La Niña and neutral periods (Table 2, Figure 5D–F). Using GLMs, associations between minimum temperature and malaria incidence were positive when stratifying by ENSO period and strongest during neutral periods. The incidence ratio for a 1°C increase in minimum temperature was 1.34 (95% CI 1.29–1.40) during neutral periods, 1.28 (95% CI 1.21–1.36) during El Niño periods (interaction *p* = 0.217), and 1.03 (95% CI 0.98–1.09) during La Niña periods (interaction *p* < 0.001) (Table 2).

**Table 2.**
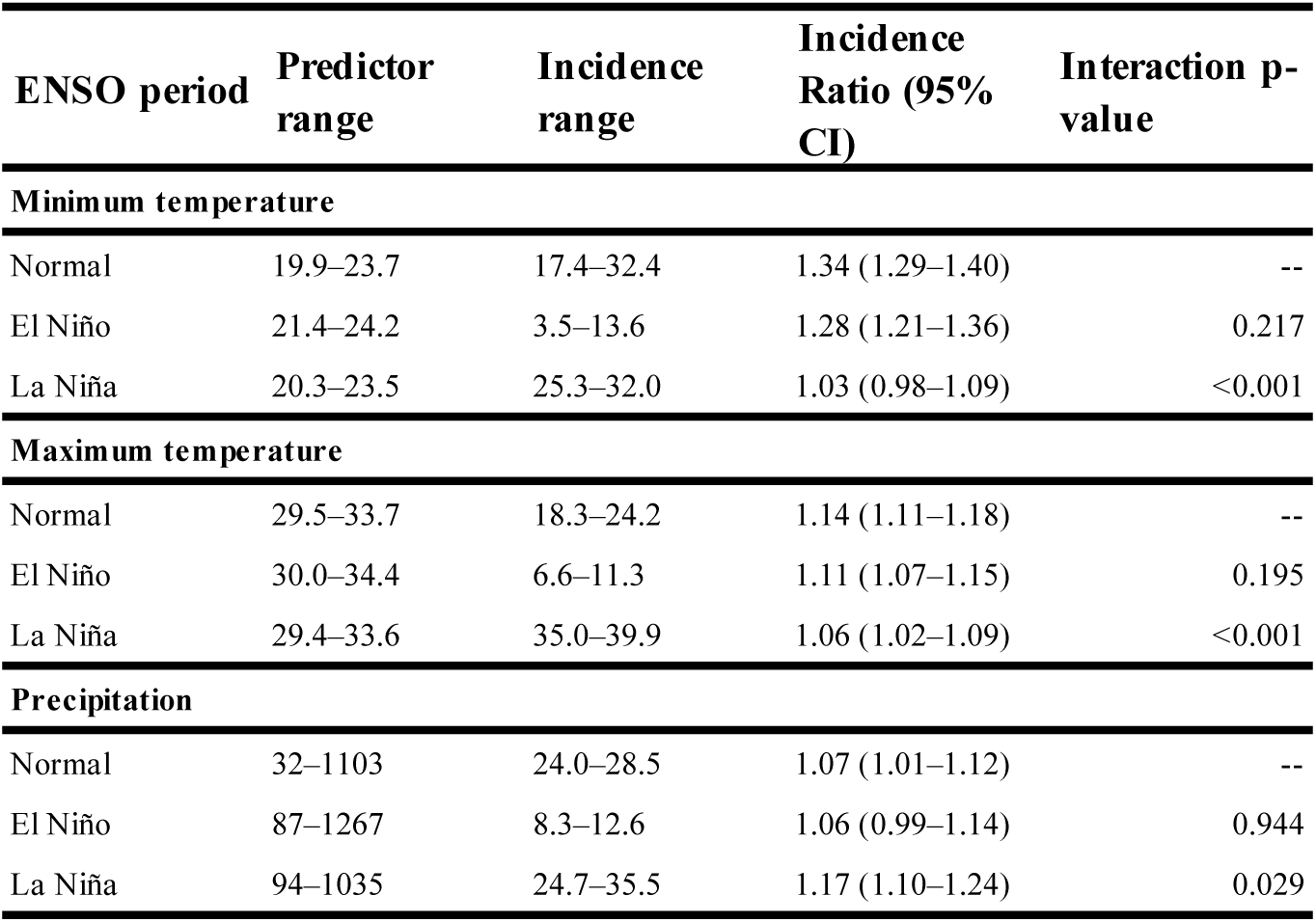

| ENSO period | Predictor range | Incidence range | Incidence Ratio (95% CI) | Interaction p-value |
| --- | --- | --- | --- | --- |
| **Minimum temperature** |  |  |  |  |
| Normal | 19.9–23.7 | 17.4–32.4 | 1.34 (1.29–1.40) | -- |
| El Niño | 21.4–24.2 | 3.5–13.6 | 1.28 (1.21–1.36) | 0.217 |
| La Niña | 20.3–23.5 | 25.3–32.0 | 1.03 (0.98–1.09) | <0.001 |
| **Maximum temperature** |  |  |  |  |
| Normal | 29.5–33.7 | 18.3–24.2 | 1.14 (1.11–1.18) | -- |
| El Niño | 30.0–34.4 | 6.6–11.3 | 1.11 (1.07–1.15) | 0.195 |
| La Niña | 29.4–33.6 | 35.0–39.9 | 1.06 (1.02–1.09) | <0.001 |
| **Precipitation** |  |  |  |  |
| Normal | 32–1103 | 24.0–28.5 | 1.07 (1.01–1.12) | -- |
| El Niño | 87–1267 | 8.3–12.6 | 1.06 (0.99–1.14) | 0.944 |
| La Niña | 94–1035 | 24.7–35.5 | 1.17 (1.10–1.24) | 0.029 |

Associations between maximum temperature and malaria incidence were positive when stratifying by ENSO period and strongest during neutral periods. The incidence ratio for a 1°C increase in maximum temperature was 1.14 (95% CI 1.11–1.18) during neutral periods, 1.11 (95% CI 1.07–1.15) during El Niño periods (interaction *p* = 0.195), and 1.06 (95% CI 1.02–1.09) during La Niña periods (interaction *p* < 0.001) (Table 2).

Associations between total precipitation and malaria incidence were positive when stratifying by ENSO period but strongest during La Niña periods. The incidence ratio for a 500mm increase in precipitation was 1.07 (95% CI 1.01–1.12) during neutral periods, 1.06 (95% CI 0.99–1.14) during El Niño periods (interaction *p* = 0.944), and 1.17 (95% CI 1.10–1.24) during La Niña periods (interaction *p* = 0.029) (Table 2).

## DISCUSSION

Higher temperatures were associated with modestly higher *Pv* malaria incidence, and total precipitation was associated with moderately higher incidence in an elimination setting in Peru. Associations generally lasted several weeks, with more sustained, stronger associations for precipitation. We only observed associations between malaria incidence and minimum temperatures above 22°C, suggesting that cooler minimum temperatures may be a limiting factor for *Pv* malaria incidence. Incidence was lowest during El Niño periods, which are typically dry in this region, and higher in wetter La Niña periods. The strength of associations was stronger during neutral periods for temperature predictors and stronger during La Niña periods for precipitation. Overall, periods of higher temperatures and cumulative rainfall were associated with increased malaria burden in the subsequent several weeks.

Our findings suggest that weather influences malaria transmission over long, 1–4-month lags in this region. This lasting association may reflect both initial infections and the relapse periodicity of *Pv* malaria. The efficacy of standard of care anti-relapse treatment is approximately 75-80%,^38^ but effectiveness can be significantly lower due to challenges with adherence.^39,40^ Further, many infections go undetected and untreated due to healthcare access challenges or lack of or minimal symptoms. As such, the vast majority of *Pv* infections are estimated to be relapse infections,^41,42^ and incidence measured in this study included both new and relapse infections. Additionally, relapses from the tropical strains of *Pv* are estimated to range anywhere from 2 weeks^4^ to 2–9 months,^21^ supporting our finding. The extended association with weather over many weeks could also reflect a high prevalence of asymptomatic and low parasitemia infections^4,5^, which could sustain new, onward transmission well past the initial meteorological trigger. Another study in this region found that PCR-confirmed *Pv* prevalence was as high as 25%, most of which were attributed to asymptomatic, low parasitemia infections.^43^

### Precipitation

We found that higher precipitation was significantly associated with higher malaria incidence at lags of 5–16 weeks. These results are generally in accordance with other studies of the effect of rainfall on *Pv* infections. A meta-analysis in Mauritania found that *Pv* incidence was highest during and after the rainy season^44^ and that decreased rainfall was significantly correlated with decreased malaria burden; a study in South Korea, a temperate area, found that increased precipitation was associated with higher malaria incidence at a 10-week lag,^45^ while a similar study in a tropical area of China also found positive associations with precipitation at lags 2–4 weeks.^46^ The lasting influence of precipitation likely reflects increased breeding ground following rainfall, which could impact multiple transmission cycles. The 5–16 week lag may also include the transition to the dry season, when the vector can still thrive in moist environments but flying and biting behavior is uninhibited by falling rain.

### Temperature

We found that higher minimum temperatures were associated with higher malaria incidence from 4–11-week lags. This finding has mixed support in the literature: though the thermal optimum of the *Pv*–*Ny. darlingi* coupling has not been studied directly, *Pf* malaria transmission peaks at 25°C,^24,47^ and minimum temperatures in our study were in the range of 20–24°C. Our finding of the strongest associations with minimum temperature is also biologically plausible given that *Ny. darlingi* typically bite humans at nighttime,^48^ and daily temperatures are lowest at night. However, a study of *Pv-*dominant Amazon border regions found that *Pv* incidence was negatively associated with minimum temperatures between 17–25°C at 0- and 6-month lags, with the stronger association at 6 months^49^. We attribute the difference at 0 months to divergent model structures and lookback periods. We also found a negative association at 16 weeks, suggesting there may be negative associations at longer lags that we did not explore due to our focus on the faster time to relapse common in our study site.

Our finding that higher maximum temperatures in the range of 30–34°C were associated with increased incidence was surprising because it conflicts with findings in neighboring Brazil^28^ and some previous disease ecology studies. One simple explanation is that during hotter temperatures, people may spend more time outdoors, or spend less time under bednets or in screened areas. However, this explanation does not address the well-documented thermal limits on vector behavior. Laboratory studies of *Pf* malaria have found that the thermal optimum for *Nyssorhynchus* is at 25°C with an upper limit of 32.6°C,^24^ and empirical studies have found that *Pf* malaria transmission declines above 28°C^47^ in Africa. However, a study in *Pv*-dominant neighboring regions of the Amazon with similar maximum temperatures (26.8–35.2°C) found that a 1°C increase in maximum temperature was associated with higher *Pv* incidence at 1- and 2-month lags, consistent with our findings.^50^ It is generally thought that *Anopheles gambiae* and *Anopheles stephensi* biting rate, larva survival, fecundity, adult survival decline at the maximum temperature range in our study (30–34°C)^25^, and a laboratory study of *Nyssorhynchus darlingi* in Brazil found that larval development, adult lifespan, and body size all declined as temperatures increased from 20°C to 28°C though there was some geographic variation.^51^ It is possible that microclimates, topographical variation, and forest cover tempered high temperatures in our study site, and such fine scale phenomena would not have been accounted for given that we used temperature data with 11km resolution. Alternatively, the vector in our study sites may be adapted to higher maximum temperatures,^52^ preventing population decline.

Another possible explanation is that associations with maximum temperature reflect relapsing *Pv* infections, which could be triggered by temperature itself, physiological conditions, or co-infections that are more common at higher temperatures in this setting. Extrinsic triggers of *Pv* relapse may include untreated *P. falciparum*^53^ or other co-infections that result in host inflammation, subsequent primary *Pv* infections, seasonal changes in sunlight and temperature, and mosquito bites and their associated immune responses.^26^ In our study site, *P. falciparum* is uncommon, though dengue is increasingly common,^54^ and it is well-documented that dengue transmission is positively associated with maximum temperatures similar to those in our study^55–57^ and has a higher thermal optimum (29°C) and upper limit (34.5°C) than malaria.^24^ Co-infections with dengue could thus occur at higher temperatures and lead to relapsing *Pv* infections. It is also possible that higher temperatures lead to host inflammation and heat stress that activate the hypnozoite and increase the reservoir of active infectious hosts, potentially leading to onward transmission and multiple cycles of infection, explaining the delayed positive association.

### ENSO sub-group analyses

We found that incidence was generally lowest during El Niño periods compared to neutral or La Niña periods, when incidence was similar. There is mixed support for this finding in the literature. One study in neighboring Colombia found that caseloads were higher during El Niño events^58^, possibly because higher temperatures and lower rainfall lead to decreased river discharge and water stagnation, creating an optimal malaria breeding ground. A study in the Brazilian Amazon found that incidence was lower during El Niño and La Niña periods^28^, somewhat contradicting our results. The rivers in our study region tend to be shallow and small, and some dry up completely during strong El Niño periods, such as in August 2023. Additionally, the Amazon has been undergoing historic drought due to global climate change^16^, with 2023 being a particularly pronounced drought year. Climate change may be dramatically altering the effects of weather independently and on a longer time scale than any individual ENSO period studied here. Drought brought on by El Niño and climate change may decrease the availability of stagnant pools of water necessary for mosquito breeding, explaining the overall lowest incidence during El Niño periods.

Additionally, temperatures were highest during El Niño periods, potentially limiting mosquito activity. On the other hand, because La Niña periods are associated with wetter conditions,^59^ in the context of historic drought that has dried up rivers in our study region, increases in rainfall could potentially create temporary pools of stagnant water, explaining the higher incidence during these periods in our study region. We also found evidence of interaction between La Niña periods and weather predictors; associations between malaria incidence and precipitation were strongest during La Niña periods, while associations between malaria incidence and temperature were weakest during La Niña periods. These interaction effects suggest that variation in precipitation drives malaria transmission during La Niña periods, while during normal and El Niño periods, temperature largely drives transmission.

### Community type sub-group analysis

Incidence was higher in river-versus highway-adjacent communities, and the association with maximum temperature was also stronger in riverine communities. On the other hand, the association with precipitation was stronger in highway-adjacent communities. It is possible that rainfall is more likely to create stagnant pools of water in highway-adjacent communities than riverine communities, where water bodies are larger and rainfall may result in a more modest increase in ideal water bodies for mosquito breeding. Conversely, in river-adjacent communities where mosquitoes can breed uninhibited, temperature may be more limiting, explaining the stronger association with maximum temperature in these communities. In addition, higher vegetation near rivers may create cooler microclimates that approximate the malaria thermal optimum better than more urbanized highway communities, which may be hotter.

### Limitations

Our study had several limitations. We were unable to distinguish between initial and relapse infections in our incidence data; we would expect that weather would influence initial and relapse cases over different lag periods, but our analysis was not able to investigate this. However, using distributed lag models allowed us to investigate associations over a 4-month period, including potential relapse cases. We were also unable to identify which cases were identified via passive surveillance versus RACD/ACD, which were done as malaria control efforts by the Peruvian Ministry of Health, greatly affecting case detection and potentially altering our results. However, these were done as a reaction to higher caseloads, suggesting that passive surveillance-detected incidence was already higher when case detection was done. A follow-up study using genomic methods to differentiate relapse and initial infections could elucidate how weather influences primary versus relapse infections. The study period also included two years (2021 and early 2022) where control efforts for the COVID-19 pandemic likely limited malaria transmission as well.

While we selected ERA-5 Land remote sensing data for its temporal coverage, its 11 km spatial resolution prevented identification of small-scale microclimates, such as those created by forest cover and topographical features, and thus did not capture small-resolution variation that may have a large effect on mosquito breeding and survival habitat.^60^ Additionally, publicly available surface water data did not reflect our ground observations of surface water in the study site, so we did not include it in this analysis. Further research that considers mediation by surface water help determine the complicated dynamics between surface water, drought, and malaria incidence in this region. While we initially considered land cover variables, they were limited in statistical power. Further research could confirm relationships between land cover, deforestation, and malaria incidence in this region and how that is mediated by climate change and drought. This study was also correlational, and inferred relationships with individual weather variables may be confounded by collinearity and correlation with other weather variables and thus made it difficult to tease out individual direct effects.

## Conclusions

In our study of malaria incidence in Amazonian Peru, we observed generally positive associations with higher temperatures and higher rainfall for extended lag periods beginning 4–5 weeks after the initial weather event and enduring for 1–4 months. We also found lower incidence during El Niño years and higher incidence in riverine communities, which may be used to concentrate resources and time malaria interventions to achieve elimination in this setting. Our findings indicated that the coupled transmission and relapse cycle of *Pv–Ny. darlingi* may have more complicated associations with higher temperatures than other malaria parasite – vector pairings, a critical finding and target for future research in the face of climate change and global warming. These findings provide critical context to ongoing malaria elimination efforts in the Amazon region, since apparent successes or failures of malaria interventions may be due in part to long-lasting effects of weather on initial infection and relapse.

## Data Availability

All data produced in the present study are available upon reasonable request to the authors.

https://osf.io/fgr6w

## Notes

### Competing Interest Statement

The authors have declared no competing interest.

### Funding Statement

This study was funded by the National Institute of Allergy and Infectious Diseases under Award Number U01AI157962.

### Author Declarations

This study was approved by the Institutional Review Board of Stanford University (72291) and by the Direccion Universitaria de Asuntos Regulatorios de la Investigacion de la Universidad Peruana Cayetano Heredia (211747).

### Summary of Updates

added additional data (2017-2020); conducted additional climate analyses; adjusted for malaria control efforts by the Peruvian government

