## Supplemental Table 1 for "Associations between weather and *Plasmodium vivax* malaria in an Amazonian elimination setting: a distributed lag analysis from 2017–2024"

**Table S1. Deviations from the pre-analysis plan.**

| **Change type** | **Predictor** | **Justification** |
| --- | --- | --- |
| Addition | Minimum temperature | Better capture overnight temperature |
| Addition | Maximum temperature | Better capture extreme heat events |
| Removed | Diurnal temperature range → daily temperature range | Simplification of analysis; temperature already covered by existing predictors (minimum and maximum) |
| Removed | Hours of low cloud cover | Lack of literature support |
| Removed | Distance to surface water | Lack of literature support |
| Removed | Distance to water flow accumulation cell | Lack of literature support, simplification of analysis, lack of statistical power to detect meaningful relationships |
| Removed | Height above nearest drainage | Simplification of analysis, lack of literature support |
| Removed | Categorical distance to forest edge | Lack of statistical power to detect meaningful relationships |
| Removed | Sub-group analysis: Above/below median distance to forest edge | Elucidate temperature & precipitation interaction effects |
| Removed | Proportion of land cover class | Lack of statistical power to detect meaningful relationships |
| Addition | Sub-group analysis: Community type | Elucidate binary predictor relationships in line with concentration-dilution hypotheses. |
| Addition | Sub-group analysis:  ENSO | Elucidate relationships in line with larger climatic cycles in the region |
